# Genetic Variation and Regulation of MICA Alters Natural Killer Cell-Mediated Immunosurveillance in Early-Onset Colorectal Cancer

**DOI:** 10.1101/2024.09.22.24314127

**Authors:** Heather M. McGee, Joseph D. Bonner, Colt Egelston, Yubo Fu, Oscar Colunga Flores, Sidney Lindsey, Lawrence Shaktah, Ferran Moratalla-Navarro, Yasmin Kamal, Kevin Tsang, Christopher P. Walker, Gregory Idos, Kevin J. McDonnell, Hedy Rennert, Elizabeth L Barry, Hermann Brenner, Daniel D. Buchanan, Peter T. Campbell, Andrew T. Chan, Jenny Chang-Claude, Jane C. Figueiredo, Manuela Gago-Dominguez, Michael Hoffmeister, Li Hsu, Jeroen R. Huyghe, Mark A. Jenkins, Loic Le Marchand, Heinz-Josef Lenz, Li Li, Annika Lindblom, Yun Ru Liu (Ruby), Brigid M. Lynch, Christina C. Newton, Kenneth Offit, Shuji Ogino, Rebeca Sanz Pamplona, Andrew J. Pellatt, Paul D. P. Pharoah, Amanda Phipps, Lorena Reynaga, Allyson Templeton, Caroline Y. Um, Alicja Wolk, Michael O. Woods, Anna H. Wu, Yen Yun, Wei Zheng, Terence M. Williams, David V Conti, Ulrike Peters, Flavio Lejbkowicz, Joel K. Greenson, Stephanie L. Schmit, William J Gauderman, Stanley R. Hamilton, Victor Moreno, Gad Rennert, Stephen B. Gruber

**Affiliations:** Department of Radiation Oncology, City of Hope National Medical Center, Duarte, CA; Department of Immuno-Oncology, City of Hope National Medical Center, Duarte, CA; Department of Medical Oncology and Center for Precision Medicine, City of Hope National Medical Center, Duarte, CA; Division of Biostatistics, Population and Public Health Sciences, University of Southern California Norris Comprehensive Cancer Center, Los Angeles, CA, USA; Oncology Data Analytics Program (ODAP), Catalan Institute of Oncology (1), L’Hospitalet del Llobregat, 08908 Barcelona, Spain; ONCOBELL Program, Bellvitge Biomedical Research Institute (IDIBELL), L’Hospitalet de Llobregat, 08908 Barcelona, Spain; Department of Clinical Sciences, Faculty of Medicine, University of Barcelona, 08007 Barcelona, Spain; Department of Medicine, Brigham & Women’s Hospital, Boston, MA; CHS National Israeli Cancer Control Center, Carmel Medical Center, Haifa, Israel; Department of Epidemiology, Department of Community and Family Medicine, Dartmouth College, Dartmouth, NH; Division of Clinical Epidemiology and Aging Research, German Cancer Research Center (DKFZ), Heidelberg, Germany; Colorectal Oncogenomics Group, Department of Clinical Pathology, Melbourne Medical School, The University of Melbourne, Parkville, Australia; Department of Epidemiology and Population Health, Albert Einstein College of Medicine, Bronx, NY, USA; Division of Gastroenterology, Massachusetts General Hospital and Harvard Medical School, Boston, Massachusetts, USA; Division of Cancer Epidemiology, German Cancer Research Center (DKFZ), Heidelberg, Germany; Department of Medicine, Samuel Oschin Comprehensive Cancer Institute, Cedars-Sinai Medical Center, Los Angeles, CA, USA; Department of Preventive Medicine, Keck School of Medicine, University of Southern California, Los Angeles, CA, USA; Public Health Sciences Division, Department of Biostatistics, University of Washington, Seattle, WA, USA Fred Hutchinson Cancer Center, Seattle, WA, USA; Public Health Sciences Division, Fred Hutchinson Cancer Center, Seattle, WA, USA; Centre for Epidemiology and Biostatistics, Melbourne School of Population and Global Health, The University of Melbourne, Melbourne, Victoria, Australia; Epidemiology Program, University of Hawaii Cancer Center, Honolulu, HI, USA; Norris Comprehensive Cancer Center, Keck School of Medicine of University of Southern California, Los Angeles, CA, USA; School of Medicine, Population Health, Cancer Control and Population Health, UVA Cancer Center University of Virginia, Charlottesville, VA, USA; Department of Clinical Genetics at Karolinska University Hospital, Stockholm, Sweden; TMU Research Center of Cancer Translational Medicine, Taipei Medical University, Taipei, Taiwan; Cancer Epidemiology Division, Cancer Council Victoria, Melbourne, Victoria, Australia; Population Science Department, American Cancer Society, Atlanta, GA, USA; Clinical Genetics Service, Inherited Cancer Genomics, Cancer Biology and Genetics, Memorial Sloan Kettering Cancer Center, Weill Cornell Medical College, New York, NY, USA; Program in MPE Molecular Pathological Epidemiology, Department of Pathology, Brigham and Women’s Hospital, Harvard Medical School, Boston, MA, USA; Department of Cancer Medicine, University of Texas MD Anderson Cancer Center, Houston, TX, USA; Cancer Epidemiology, Department of Public Health and Primary Care, Cambridge Cancer Centre University of Cambridge, Cambridge, UK; Department of Epidemiology, University of Washington, Seattle, WA, USA; Institute of Environmental Medicine, Karolinska Institutet, Stockholm, Sweden; Memorial University of Newfoundland, St. John’s, Newfoundland, Canada; Department of Population and Public Health Sciences, University of Southern California, Los Angeles, CA, USA; Cancer Biology and Drug Discovery, Taipei Medical University, Taipei, Taiwan; Division of Epidemiology, Vanderbilt Epidemiology Center, Vanderbilt-Ingram Cancer Center, Nashville, TN, USA; Public Health Sciences Division, Fred Hutchinson Cancer Center, Department of Epidemiology, University of Washington, Seattle, WA, USA; Department of Pathology, University of Michigan, Ann Arbor, MI, USA; Genomic Medicine Institute, Lerner Research Institute, Cleveland Clinic, Cleveland, OH, USA; Department of Pathology, City of Hope National Medical Center, Duarte, CA, USA; B Rappaport Faculty of Medicine, Technion-Israel Institute of Technology, Technion and the Association for Promotion of Research in Precision Medicine (APRPM), Haifa, Israel

## Abstract

The incidence of colorectal cancer (CRC) among individuals under age 50, or early-onset CRC (EOCRC), has been rising over the past few decades for unclear reasons, and the etiology of the disease remains largely unknown. Known genetic risk factors do not explain this increase, pointing to possible environmental and as-yet unidentified genetic contributors and their interactions. Previous research linked genetic variation on chromosome 6 to increased CRC risk. This region harbors multiple immune genes, including the gene encoding Major Histocompatibility Complex (MHC) class I polypeptide-related sequence A (MICA). MICA is a polygenic ligand for the Natural Killer Group 2D receptor (NKG2D), a receptor expressed on Natural Killer (NK) cells and other lymphocytes. Given that intra-tumoral NK cell infiltration correlates with favorable CRC outcomes, we hypothesized that germline genetic variation in *MICA* could influence CRC risk. In a discovery set of 40,125 cases and controls, we show that the minor G allele at Chr6:31373718C>G (hg19) is associated with increased risk for CRC (odds ratio (OR) = 1.09, 95% confidence interval (CI) 1.04 - 1.15, p = 0.0009). The effect is stronger in EOCRC (OR = 1.26, 95% CI 1.08 - 1.44, p = 0.0023) than in those 50 and over (OR = 1.07, 95% CI 1.02 - 1.13; p = 0.012) (Ratio of ORs = 1.32, 95% CI 1.14 - 1.52, p = 0.0002). In an independent validation set of 77,983 cases and controls, the adjusted interaction by age-of-onset was significant at OR = 1.15 (95% CI 1.03 - 1.34, p = 0.0150) with a higher risk in EOCRC. Expression quantitative trait locus analysis in normal colonic epithelia showed that MICA RNA expression decreases linearly with each additional copy of the minor G allele (p = 3.345 × 10e-18). Bulk RNA analysis of the tumor immune microenvironment revealed that tumors from patients with CG or GG genotypes have lower resting and activated NK cell infiltration as compared to tumors from patients with CC genotype. Multiplex immunofluorescence analysis demonstrated that patients with a G allele (i.e. CG or GG genotype, but not CC genotype) have a statistically significant decrease in the number of NK cells in tumor compared to adjacent normal colonic mucosa. Taken together, population-based epidemiologic, molecular, genetic, cellular and immunologic evidence demonstrate that *MICA* genotype is associated with increased risk of EOCRC and reduced number of NK cells in colorectal tumors, suggesting that patients with a G allele have altered NK cell-mediated immunosurveillance. These novel findings suggest that EOCRC may have a previously unrecognized innate immune-mediated etiology which merits further investigation.

## Background

### MHC class I polypeptide-related sequence A (MICA)

MHC class I polypeptide-related sequence A (MICA) encodes a membrane-bound protein that serves as a stress-induced ligand. Most healthy cells do not express MICA, except for intestinal epithelial cells (*2*), endothelial cells and fibroblasts (*3*). MICA expression is upregulated in response to stressors such as tissue damage, infection, cellular transformation, environmental exposures, and therapeutic agents, including vitamins C and E, thermal stress, chemotherapy, and ionizing radiation (*4-9*). When cells express MICA on their surface, MICA binds to the C-type lectin natural killer group 2D (NKG2D; official symbol KLRK1) receptor on NK cells, γδ T cells, CD8+ T cells, NK-T cells and innate lymphoid cells (ILCs), leading to activation of these immune cell subsets. Viruses such as Cytomegalovirus (CMV), Human Immunodeficiency Virus (*10*), and Hepatitis B Virus (HBV) have evolved mechanisms to manipulate MICA expression and evade the immune response. Similarly, certain cancers such as melanoma, lung, triple-negative breast, and colon cancer can decrease MICA expression or release MICA in its soluble form to escape immune surveillance (*11-14*).

### Colorectal Cancer Epidemiology

Colorectal cancer (CRC) is the third leading cause of cancer-related death (*15*). One of the most striking and unexplained recent trends in oncology is the sharp rise in early-onset colorectal cancer (EOCRC) in patients under 50 years old since the mid-1990s (*16, 17*). EOCRC now accounts for 10% of all new CRC diagnoses (*18*) and is the leading cause of cancer-related death among men under 50 and the second leading cause among women under 50 (*19*). Younger patients often present with more advanced disease, frequently exhibiting poorly differentiated tumors with signet-ring cell features, which are associated with a worse prognosis (*20, 21*).

EOCRC can be observed in patients with genetic predisposition syndromes, including Lynch Syndrome (*MLH1, MSH2, MSH6, PMS2, EPCAM*), Familial Adenomatous Polyposis (*APC*), Li-Fraumeni syndrome (*TP53*), MUTYH-associated polyposis (*MUTYH*), Cowden syndrome (*PTEN*), and Peutz-Jeghers syndrome (*STK11*) (*22*). However, these syndromes do not fully explain the rising incidence of EOCRC. The rate of increase in EOCRC incidence outpaces the stable population frequency of these well-characterized mutations and syndromes, indicating that other factors are likely at play (*23*).

### Immune Cells Involved in Colorectal Cancer

The immune system plays a role in both the development and prognosis of colorectal cancer. Infiltration of CD8+ T cells in CRC is associated with a favorable prognosis (*24-27*) since T cells are the cornerstone of the adaptive immune system, defined by their antigen specificity and their ability to form immunologic memory. Recent work suggests that the diversity, rather than the abundance, of T cells, may influence EOCRC risk (*1*).

Human chromosome 6 encodes nearly 200 genes within the Major Histocompatibility Complex (MHC). This highly polymorphic region contains Human Leukocyte Antigen (HLA) genes that encode the α chains of MHC class I molecules and the α and β chains of MHC class II molecules, as well as Major Histocompatibility Complex Chain I polypeptide-related sequence A and B (MICA/ MICB), Tumor necrosis factor (TNF) and Nuclear factor kappa beta inhibitor like 1 (NFKBIL1). MHC molecules bind to antigen-derived peptides, and the MHC-peptide interaction with the T cell receptor (TCR) leads to T cell activation. Significant research has focused on how HLA abundance and diversity affects the adaptive immune response (*28*). Our recent studies report that genomic variation within chromosome 6 contributes to CRC risk and prognosis (*29-32*).

There is also evidence linking CRC outcomes to Natural Killer (NK) cells, innate lymphocytes that lack antigen specificity. Low pre-operative NK cell cytotoxicity significantly predicted CRC recurrence after surgery, CRC metastasis, development of metachronous disease and 5-year survival (*10, 33, 34*). NK cell abundance in the peripheral blood was associated with a modest reduction in cancer risk (*35*). Our group has reported that increased RNA expression of NK cell markers is associated with favorable CRC-specific survival and progression-free survival (*36*). Interestingly, NK cells are often abundant in normal mucosal tissue but scarce in adjacent abnormal colorectal cancer (*37*).

NK cells are activated by either downregulation of Major Histocompatibility Complex I (MHC I) or through signaling via activating and inhibitory receptors. The most well-studied NK cell activating receptor is NKG2D, which is encoded by the Killer cell lectin like receptor K1 (KLRK1) gene. In humans, NKG2D binds to a group of stress ligands, called NKG2D ligands (NKG2DL), which include Major Histocompatibility Complex Chain I polypeptide-related sequence A and B (MICA/ MICB) and Unique Long 16 (UL16) binding proteins (ULBP-16). Genomic variation in NKG2D (on chromosome 12) may affect lifetime CRC risk (*38*) and CRC progression (*33, 39*).

### Natural Killer (NK) cells in Viral Infection

Some human viral infections, such as CMV, Hepatitis B, and HIV, interfere with NK cell biology. CMV can sequester MICA, preventing it from reaching the cell surface and activating NK cells (*40*). Hepatitis B downregulates MICA expression in infected liver cells, hampering NK cell recognition (*41*). HIV is particularly adept at downregulating MICA so the virus can evade NK cell cytotoxicity (*42*). Some individuals can control the progression of HIV infection over time. The genetics of these HIV long-term non-progressors have been studied extensively. In their 2014 study, LeClerc et al. analyzed imputed genotype values from a genome-wide association study (GWAS) to identify genotypes associated with non-progression and elite response (*43*). They reported the strongest associations for non-progression on chromosome 6, between positions 30993567 and 31431780. They identified several SNPs in this region that are more common in non-progressors, suggesting that germline variation in this region may confer protection from HIV infection through an immune pathway involving MICA.

We hypothesized that genotypes in and around *MICA* modulate NK cells in the colon to alter immunosurveillance and contribute to the development of EOCRC. To investigate our hypothesis, we analyzed genotypes in the target region of chromosome 6 harboring *MICA* in a large well-characterized consortium study of CRC and validated the finding in a larger independent validation study. Next, we evaluated *MICA* expression differences in normal colonic tissue via expression quantitative trait locus analysis (eQTL). In addition, we quantified differences in the number of NK cells in colorectal cancer tissues from cases across genotypes using both RNA sequencing and multi-plex immunohistochemistry.

## Results

### Association Discovery

The discovery set for this analysis was from genotyping cases and controls in the CORECT consortium (*44*). The discovery set includes 40,125 participants, including 23,975 cases and 16,150 controls, and compiled results from 21 individual studies (**Supplemental Table 1**). Most (37,335/40,125; 92%) of cases had an age at diagnosis and most of controls had an age at cancer-free enrollment. The dataset is comprised of 47% females (53% males) and includes 2,938 cases of EOCRC (patients diagnosed before 50 years old). Demographic information about the clinical characteristics of patients in this consortium is shown in **Table 1a**.

**Table 1.**

| **Table 1a: Characteristics of the Discovery Set** | **Group** |  |  |
| --- | --- | --- | --- |
| <u>Characteristic</u> | <u>Overall</u> | <u>Case</u> | <u>Control</u> |
| N | 40,125 | 23,975 | 16,150 |
| Mean Age (*17*) | 64 (13) | 64 (12) | 64 (14) |
| Male | 21,199 | 13,230 | 7,969 |
| Female | 18,926 | 10,745 | 8,181 |
| Under 50 | 4,959 | 2,938 | 2,021 |
| 50+ | 32,376 | 20,944 | 11,432 |
| **Table 1b: Characteristics of the Validation Set** |  |  |  |
| N | 76,983 | 33,994 | 42,989 |
| Mean Age (*17*) | 62(11) | 64(10) | 61(11) |
| Male | 39,666 | 18,148 | 21,518 |
| Female | 37,317 | 15,846 | 21,471 |
| Under 50 | 8,028 | 3,057 | 4,971 |
| 50+ | 68,955 | 30,937 | 38,018 |

The CORECT genomic data contains 449 SNPs on Chromosome 6 between coordinates 31368488 – 31383092 highlighted by LeClerc et al. in and around *MICA* (*43*). Most (362/449) of the SNPs from this region were pruned for linkage disequilibrium R^2^ above 0.2. The remaining 87 (11 directly genotyped and 76 imputed) candidate SNPs were tested for association with CRC. Association tests were adjusted for age, study, genotyping platform and three principal components of global ancestry (**Figure 1**). The SNP rs9295988 (Chr6:31373718C>G within the MICA gene) is statistically significantly associated with odds of CRC (p-value of 0.00079, FDR 0.069).

**Figure 1:**
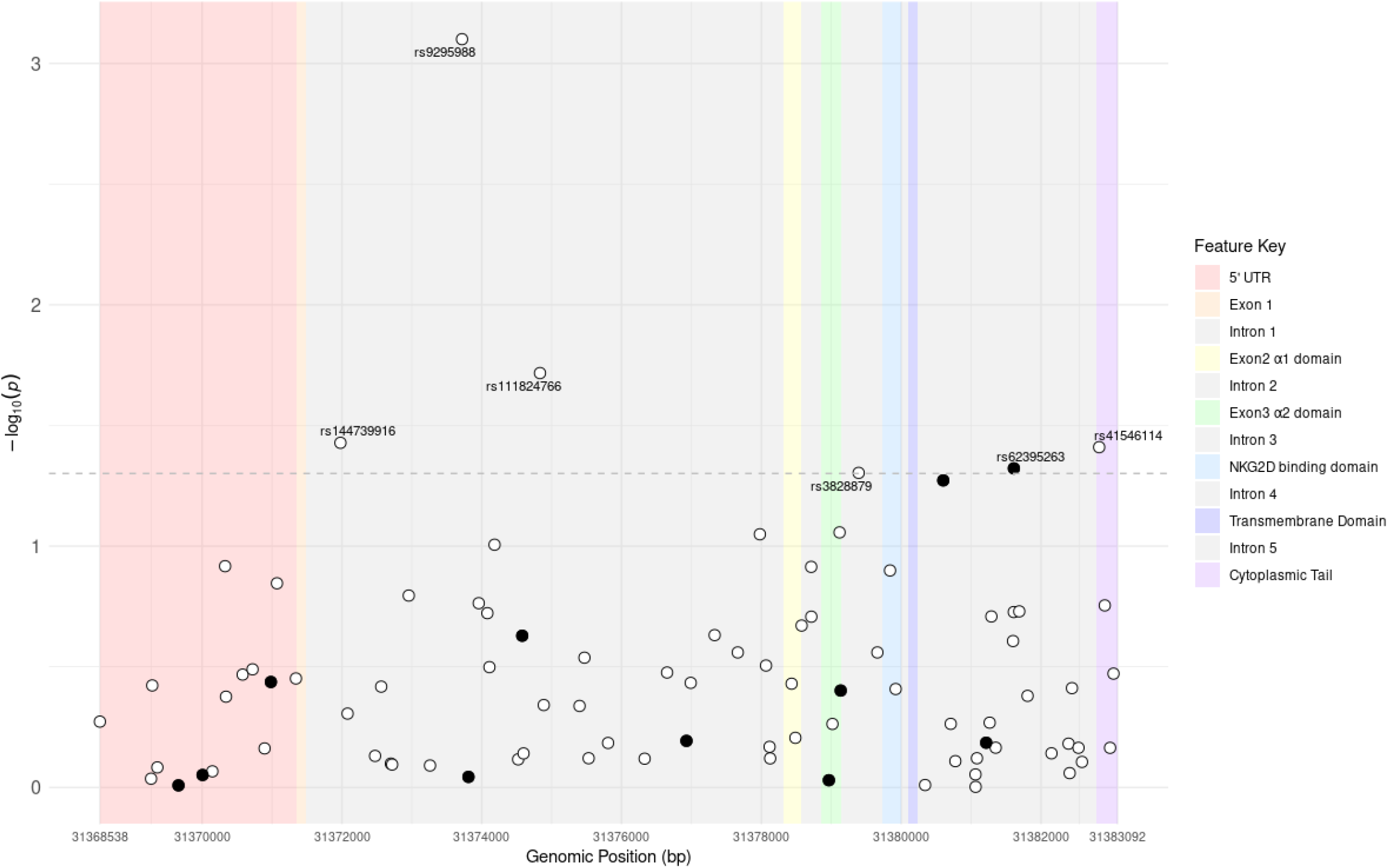
Locus zoom plot showing associations of SNPs with risk of CRC in and around MICA gene on chromosome 6. Exon/Intron boundaries are designated by color with functional domains annotated within exons. Directly genotyped SNPs are shown with solid circles, imputed SNPs with open circles. Significance threshold for association with risk of colorectal cancers shown with horizontal dotted line at nominal p-value <0.05 after adjustment for age, sex, three principal components of ancestry and study center.

The lead SNP (rs9295988) G allele is found in 20.4% of cases and 19.6% of controls of all ages (**Table 2a**) and is significantly associated with CRC overall (OR: 1.09; 95% CI: 1.04-1.15; p-value: 0.0009). This association is driven mostly by EOCRC. Among those under 50, the lead SNP is associated with a per-allele increased risk of CRC of 1.259 (95% CI: 1.083-1.442; p-value: 0.002), whereas there is little appreciable association among those over 50 (OR: 1.071, 95% CI: 1.015-1.131, p-value: 0.012). In a model fully adjusted for age, study, genotyping platform, and three principal components for ancestry, this effect is clearly stronger in EOCRC; the interaction model term from the per-allele dosage model is ROR 1.316 (95% CI: 1.139-1.522, p-value: 0.0002) demonstrating that the effect is 31% stronger in EOCRC (**Table 2a**).

### Association Validation

We validated the association with rs9295988 in an independent set of 76,983 participants (33,994 cases and 42,989 controls, **Table 1b**) from 42 studies. The demographic and clinical characteristics of participants from the validation study are presented in **Supplemental Table 2**. In the validation set, a per-allele dosage genotype model independently confirmed a stronger effect of MICA genotype in EOCRC with ROR = 1.15 (95% CI: 1.03-1.29, p-value:0.0150) (**Table 2b**).

**Table 2.**

| **Table 2a: Dosage Model Association in the <u>Discovery Set</u>** |  |  |  |  |
| --- | --- | --- | --- | --- |
|  | Cases |  | Controls |  |
| Genotype | Under 50 | 50 and over | Under 50 | 50 and over |
| C/C | 2,306 | 16,712 | 1,660 | 9,110 |
| C/G | 563 | 3,889 | 340 | 2,141 |
| G/G | 69 | 343 | 21 | 181 |
| Adjusted Interaction term: ROR: 1.315 (95%CI: 1.138-1.522, p-value:0.0002) |  |  |  |  |
| **Table 2b: Dosage Model Association in the <u>Validation Set</u>** |  |  |  |  |
|  | Cases |  | Controls |  |
| Genotype | Under 50 | 50 and over | Under 50 | 50 and over |
| CC | 2,491 | 4,146 | 25,567 | 31,668 |
| CG | 524 | 774 | 5,012 | 5,989 |
| GG | 42 | 51 | 358 | 361 |
| Adjusted Interaction Term ROR: 1.15 (95% CI: 1.03-1.29, p-value:0.0150) |  |  |  |  |

### Expression Quantitative Trait Locus (eQTL)

To assess the role of SNP rs9295988 on the expression of *MICA* RNA expression in the normal colonic epithelium, we performed expression quantitative trait locus analysis. Using the Colonomics resource, (*45*) we discovered that each additional G allele of rs9295988 decreases the expression of MICA by almost 1 standard deviation in healthy colonic epithelium. Using the BarcUVaSeq database, we found a statistically significant inverse relationship between each G allele and MICA expression, with a slope of -0.870 (SD 0.096), p = 3.3e-18 (**Figure 2**).

**Figure 2:**
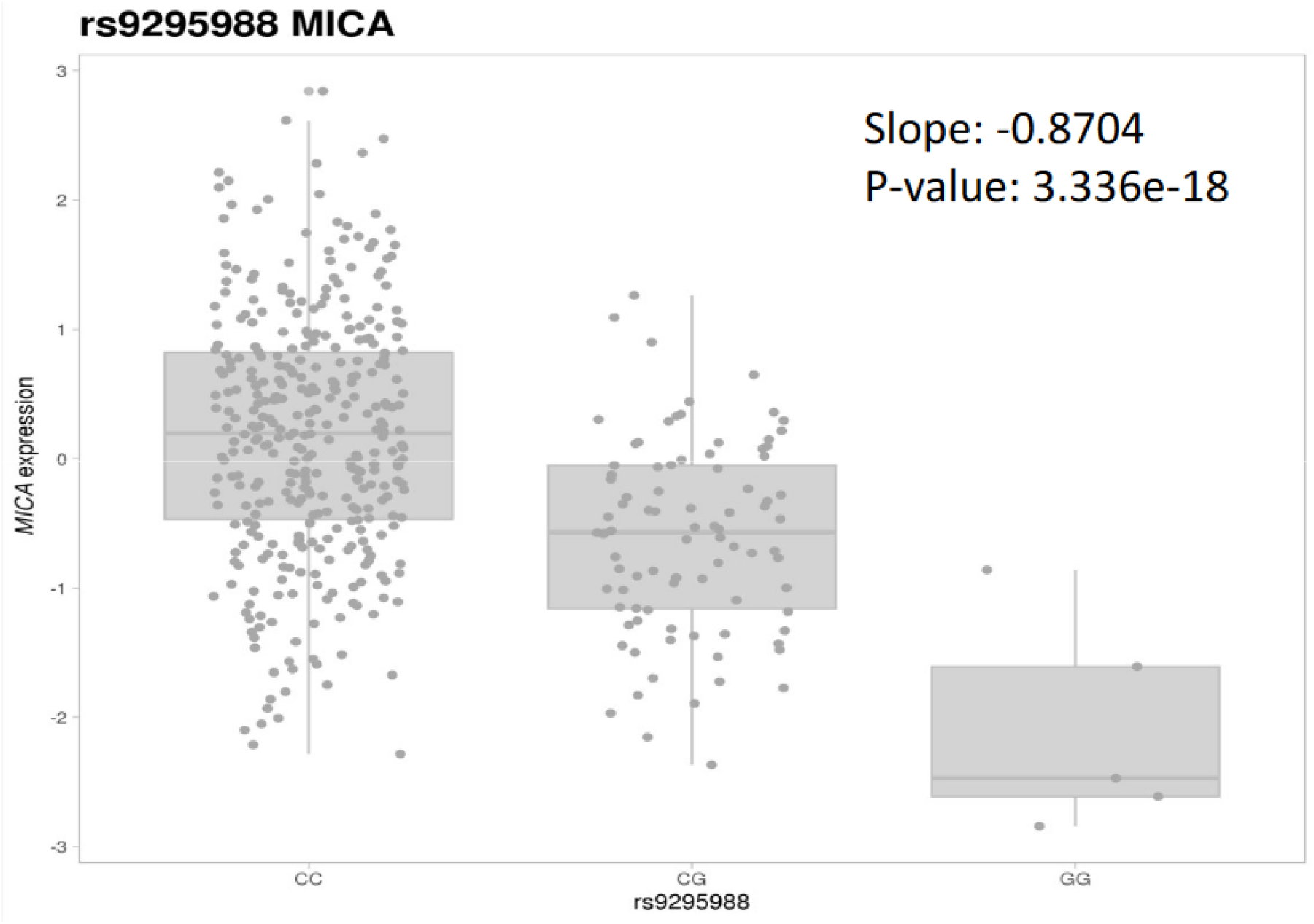
MICA RNA expression differences across rs9595988 genotypes in normal colonic epithelia. eQTL analysis of normal colonic mucosa from BarcUVaSeq (a publicly available database) demonstrates MICA expression differences by rs9295988. Trends for statistical significance analyzed by linear regression.

### Deconvoluted RNA expression analysis

To determine whether various genotypes lead to alterations in NK cells, we utilized RNA expression data from the Molecular Epidemiology of Colorectal Cancer (MECC) study, which is part of the CORECT consortium. MECC is a population-based, incidence density case-control study of incident colorectal cancer diagnosed within a geographically defined region of northern Israel between March 31, 1998 and July 1, 2017.

The BASE deconvolution algorithm (*46*) was applied to bulk tumor gene expression data from a subset of 270 MECC frozen tumor samples. The algorithm quantifies immune infiltrates from 22 different cell types using the LM22 immune cell signature matrix. We used this dataset to compare the relative expression level of markers of resting and activated NK cells in specimens from CRC patients with each genotype. This analysis revealed that NK cells in both resting and activated state are downregulated in tumors from individuals carrying a minor G allele at rs9295988 (i.e. patients with both CG and GG genotypes) (**Figure 3a and 3b**).

**Figure 3:**
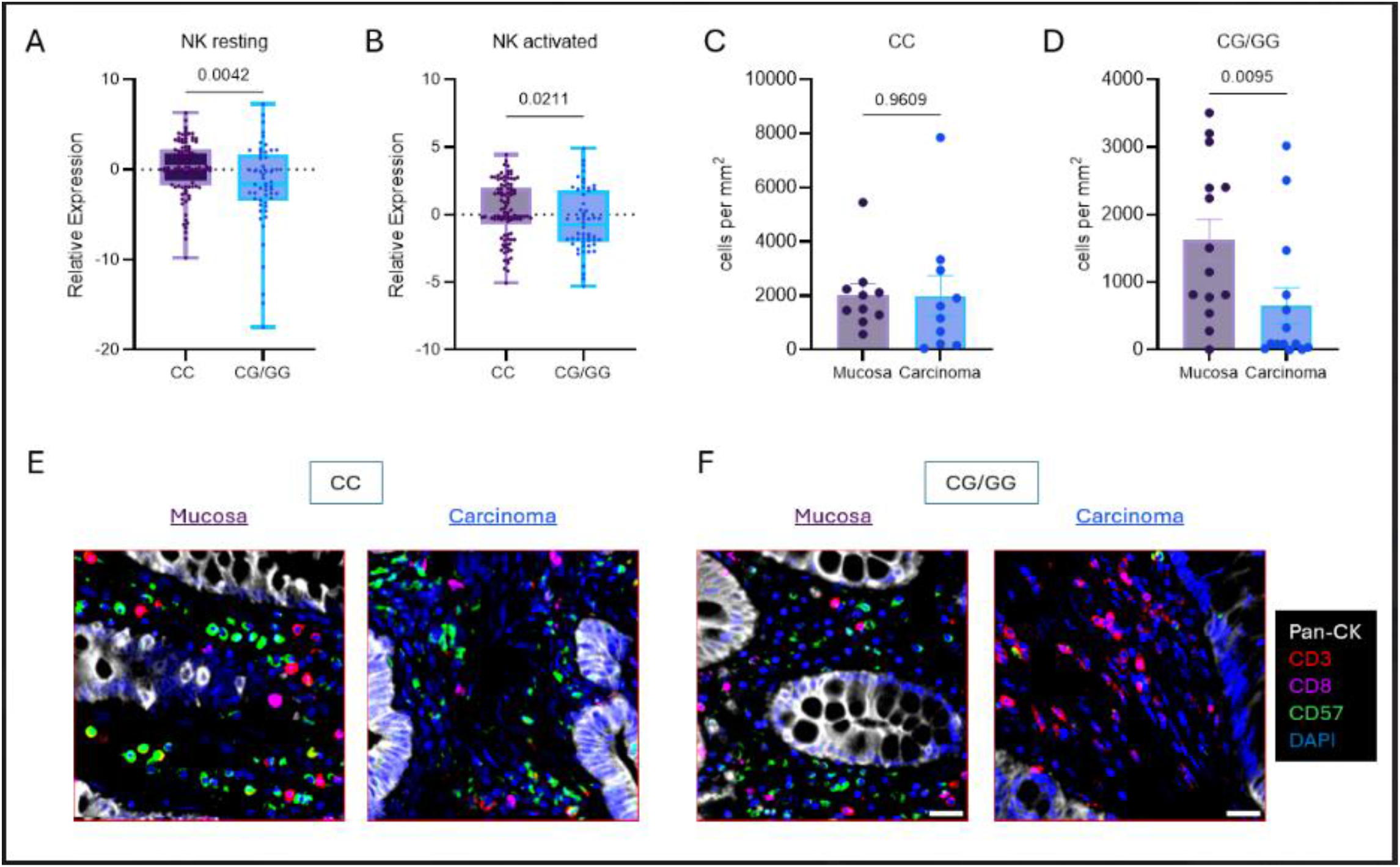
Comparison of NK Cells in Colorectal Carcinoma and Normal Colonic Mucosa by *MICA* rs9295988 Genotype. A. Relative expression of resting NK cell signature by rs9295988 genotypes from bulk RNA expression in macrodissected colorectal adenocarcinoma from snap-frozen tissue (CIBERSORT data). B. Relative expression of resting NK cell signature by rs9295988 genotypes from bulk RNA expression in macrodissected colorectal adenocarcinoma from snap-frozen tissue (CIBERSORT data). C. Quantification of the number of NK cells per mm^2 tissue in the normal colonic mucosa vs. colorectal carcinoma of patients with CC genotype. n = 10 per genotype. D. Quantification of the number of NK cells per mm^2 tissue in the normal colonic mucosa vs. colorectal carcinoma of patients with a G allele at rs9295988 (CG or GG genotype). n = 10 per genotype. E. Multiplex immunofluorescence of CD57+ NK cells, CD8+ CD3+ T cells and CD3+ CD8-CD4 T cells in normal colonic mucosa vs. colorectal carcinoma from patients with the CC genotype at rs9295988. F. Multiplex immunofluorescence of CD57+ NK cells, CD8+ CD3+ T cells and CD3+ CD8-CD4 T cells in normal colonic mucosa vs. colorectal carcinoma from patients with a G allele at rs9295988 (CG or GG genotype).

### Multiplex immunofluorescence

In order to investigate a role for NK cells at the protein level, we performed multiplex immunofluorescence to visualize and quantify the number of CD4+ T cells, CD8+ T cells, NK cells (CD57+), and NKT cells in the regions of normal colonic mucosa and tumor delineated by a board-certified Pathologist (**Supplementary Figure 1**). This analysis revealed that patients with a G allele (either CG or GG genotype) have a statistically significant decrease in the number of NK cells in tumor compared to adjacent normal colonic mucosa (student’s t-test; p = 0.0095) (**Figure 3d and 3f**), while CC carriers have a similar number of NK cells in tumor and normal colonic mucosa (ns) (**Figure 3c and 3e**). This difference was not observed for any of the other lymphocyte populations examined (CD4+ T cells, CD8+ T cells or NK-T cells) (data not shown).

## Discussion

The increasing incidence of EOCRC has prompted a global call to action for oncologists and researchers to identify its causes and develop preventive strategies (*47, 48*). Although rare genetic syndromes are more common in EOCRC as compared to average age-of-onset CRC (AOCRC), Mendelian syndromes clearly do not explain rising trends in the incidence of EOCRC. Among other potential contributors (e.g. obesity, Western diet, antibiotic exposure), evidence supports multiple roles that the immune system may play in the clinical trajectory and therapeutic responses of CRC. Here, we explore how dysregulation of the innate immune system plays a pathogenic role in the development of EOCRC.

We hypothesized that G alleles at the rs9295988 SNP in *MICA* would lead to a decrease in the number of NK cells in the colorectal microenvironment, and this would contribute to altered immunosurveillance and subsequently lead to CRC pathogenesis. While some groups have shown that an increase in NK cells is associated with a better prognosis in CRC (*49*), many large-scale analyses of immune infiltrates in CRC have failed to include NK cells (*50*). Therefore, the precise role of NK cells in CRC risk is unknown.

Our large-scale genomic analysis reveals that the *MICA* rs9295988 variant increases the risk of CRC specifically in younger adults (less than 50 years old), but this SNP has less association with CRC in patients over age 50. We also observed that carriers of the minor allele of rs9295988 exhibit decreased expression of RNA signatures of resting and activated NK cells in colorectal tumors. In addition, we found that patients with a G allele (either CG or GG genotype) have a statistically significant decrease in the number of NK cells in tumor as compared to normal colonic mucosa, while CC carriers have a similar number of NK cells in tumor versus normal colonic mucosa. This strongly suggests that patients with a G allele have altered NK cell-mediated immunosurveillance that could contribute to the development of CRC. Importantly, this is corroborated by previous studies which have shown that NK cells are abundant in normal mucosal regions but scarce in adjacent abnormal colorectal cancer (*37*).

Interestingly, MICA can be expressed as cell membrane-bound form or secreted as a soluble form. A seminal study showed that tumor-derived soluble MICA induces the endocytosis and degradation of the NKG2D receptor, causing a reduction in NKG2D expression on immune cells in the tumor microenvironment as a mechanism of tumor immune-mediated evasion (*2*). However, the mechanism by which soluble MICA is secreted is largely unknown, and our study raises the possibility that this intronic SNP may allow for enhanced secretion of soluble MICA as a means of dysregulating tumor immunosurveillance.

The cancer immunoediting hypothesis proposed by Schreiber and colleagues describes the interaction between cancer cells and the immune system in three phases (*51*). The elimination phase reflects the early recognition and elimination of cancer cells that develop within normal epithelium, to control the establishment of tumors. The second phase, equilibrium, reflects a homeostatic balance between cancer cells and the immune system. The third phase, escape phase, is characterized by immunosuppressive mechanisms that create a permissive environment to allow for tumor progression. Our study implicates MICA regulation in the pathogenesis of EOCRC and suggests that EOCRC is at least partially mediated by reduced NK cell immunosurveillance. Therefore, our novel findings suggest that NK cells may play an important role in cancer immunoediting and the prevention of EOCRC (*52*).

The timeframe during which EOCRC has increased is not consistent with the generational time required for the SNP rs9295988 minor allele frequency to have risen according to the Haldane hypothesis (*53*), so change in allele frequency of MICA genotypes over time is not likely to explain EOCRC trends. Complex interactions between germline genotype and environmental factors such as diet influence the risk of a disease in carriers. Therefore, it is likely that the immune-mediated effects of *MICA* SNP rs9295988 may be exaggerated by environmental factors. We suspect that if the minor G allele at rs9295988 increases soluble MICA, then soluble MICA could also be induced by environmental factors (such as Western diet, low fiber intake, increased antibiotic use), and this combination of factors could help explain the increase in EOCRC incidence over the past few decades.

Until now, little data has linked the etiology of EOCRC to immune mechanisms, especially within the context of the innate immune system. Our characterization of the role of MICA genotype at rs9295988 is significant and meaningful, as rs9295988 is the first genetic variant linked specifically to EOCRC. Our results further show that dysregulated NK cell-mediated immunosurveillance in normal colonic mucosa may be a key mechanism contributing to the pathogenesis of colorectal cancer in young patients <50 years old. By demonstrating that MICA is a contributing genetic determinant of EOCRC, our work contributes to a paradigm shift in which EOCRC can, in part, be viewed as an immune-mediated disease.

## Methods

### Association Discovery

Quality-controlled genotype data were analyzed using PLINK software (*54*). SNPs in linkage disequilibrium above R2>0.2 were pruned. We quantified the risk of carrying a minor allele in any SNPs in LeClerc (*43*) region encompassing MICA among CRC cases relative to CRC-free controls using a logistic regression model adjusting for principal components of genetic ancestry, age, sex, and genotyping platform.

The CORECT Study is a large, multi-ethnic international collaborative GWAS consortium that has led to the identification of hundreds of novel loci contributing to risk of developing CRC (*32, 55*). Descriptions of studies contributing to the CORECT consortium have been detailed previously (*29, 44*). Briefly, CRC cases and controls of European and Asian descent from 21 observational studies (**Supplemental Table 1**) contributed data and germline DNA (Supplemental Table 1). Genotypes were derived from three different arrays and imputed to 1000 Genomes Project Phase 1 multiethnic reference panel (March 2012 release, n=1,092); (*56*)

### Association Validation

The significant SNP was validated in an independent set of 70,983 individuals of the FIGI consortium. The genomic methods of this study have been detailed elsewhere (*57*). In short, genotypes of the GECCO (*58*) consortium for participants with environmental exposure from 41 different studies were processed and quality-controlled using the BinaryDosage package and assembled to conduct gene-environment-wide-interaction studies (GEWIS) using GxEScanR (*57*). For validation purposes in the current study, CORECT participants that were also in FIGI and were removed to assure full independence (**Supplemental Table 2**). The R package (glm function) was used to quantify the risk of carrying a minor allele in SNP rs9295988 among CRC cases relative to CRC-free controls using a logistic regression model adjusting for principal components of genetic ancestry, age, sex, and study.

### Expression Quantitative Trait Locus (eQTL)

The association between genotypes of candidate SNPs and gene expression levels within normal colonic epithelium was performed using data that are published and publicly available through Colonomics (https://www.colonomics.org/) (*45*) and BarcUVaSeq (www.barcuvaseq.org) online resources (*59*).

### Deconvoluted RNA expression analysis

Detailed RNA expression data, unstained recut slides and other variables are available from one of the studies that contributed to the CORECT consortium, the Molecular Epidemiology of Colorectal Cancer (MECC) study for this analysis. MECC is a population-based, incidence density case-control study of incident colorectal cancer diagnosed within a geographically defined region of northern Israel between March 31, 1998 and July 1, 2017. Methods have been previously described (*60, 61*).

The BASE deconvolution algorithm (*46*) was applied to bulk tumor gene expression data from a subset of 270 MECC frozen tumor samples. The algorithm quantifies infiltrates from 22 different immune cell types, including resting and activated NK cells using the LM22 immune cell signature matrix, which includes 547 genes used to define 22 immune cell subtypes. The resultant values show the relative expression level of an individual cell type.

### Multiplex Immunofluorescence

Formalin-fixed paraffin-embedded (FFPE) tissue specimens from the MECC cohort were cut at 3-4 µm sections and baked onto glass slides. The FFPE slides were then deparaffinized in xylene and then rehydrated in decreasing ethanol concentration washes. Heat-induced antigen retrieval was performed using boiling AR9 buffer, 10x (pH 9) (Akoya Biosciences) in a microwave oven for 20 minutes. Blocking was performed for 10 minutes using Antibody Diluent with Background-Reducing Components (Agilent) to minimize non-specific background staining. Primary antibodies were incubated for 1 hour on a shaker at room temperature followed by a 10-minute incubation of horseradish peroxidase (HRP)-conjugated secondary antibody (Mouse HRP-Polymer, Biocare Medical). Immunofluorescent labeling of antibodies was achieved using the OpalTM 7-color fluorescence IHC Kit (Akoya Biosciences) at a 1:100 dilution for 10 minutes. Slides were serially stained with the microwave incubation acting to remove previous antibodies while simultaneously exposing the next epitope of interest. After staining the final marker, cell nuclei were stained with DAPI (Akoya Biosciences) and the slides were mounted with ProLong Gold Antifade Reagent (ThermoFisher Scientific). Primary antibodies were as follows: CD3 (clone LN10, Leica), CD8 (clone 4B11, Leica), B3GAT1 (CD57; clone HNK-1, Biolegend), and KRT20 (cytokeratin 20; clone Ks20, Dako). Tissue slides were scanned using the Vectra 3.0 automated quantitative pathology imaging system (Akoya Biosciences). Images were spectrally unmixed with inForm® tissue analysis software (Akoya Biosciences) and component TIFFs were exported for whole slide analysis using QuPath software(*62*). Mucosa and superficial tumor areas were manually chosen with the aid of a board-certified pathologist (SH). Within tumor and mucosal areas, tissue stroma was segmented from epithelial cells using a pixel classifier trained with representative training images from every slide. Within tissue stroma, cells were segmented using a watershed cell detection approach based on a DAPI nuclear expansion. Identified cells were then classified as being positive or negative for CD3, CD8, or CD57 based on a cell classifier trained with representative training images from every slide. Cell counts and tissue areas were then exported for calculation of cell densities for identified cell phenotypes.

## Statistical Methods

Genomic analyses were conducted in PLINK (*54*). Other statistical analytical scripting was conducted in the R-platform (*63*). Mean RNA expression and NK cell quantification differences were tested for statistical significance with t-tests.

## Data Availability

All data produced in the present study are available upon request to the authors.

