## Supplemental Tables and Figure for "Genetic Variation and Regulation of MICA Alters Natural Killer Cell-Mediated Immunosurveillance in Early-Onset Colorectal Cancer"

| Supplementary Table 1. Descriptors of Discovery Set Studies | | | | | | | | | | | | | | |
|---|---|---|---|---|---|---|---|---|---|---|---|---|---|---|
| | | | | | Cases | | | | | Controls | | | | |
| | | | | | | | Genotype | | | | | Genotype | | |
| Study | Title | StudyType/ Population | Total | Females | N | <50 | GG | GC | CC | N | <50 | GG | GC | CC |
| ATBC | Alpha-Tocopherol, Beta Carotene Cancer Prevention Study | cohort/ Finland | 177 | - | 148 | - | 125 | 22 | 1 | 29 | 1 | 26 | 3 | - |
| CFR | Colon Cancer Family Registry | case-control/ USA, Canada, Australia | 4,254 | 2,078 | 2,803 | 1,184 | 2,124 | 596 | 83 | 1,451 | 631 | 1,176 | 250 | 25 |
| CPSII | American Cancer Society Cancer Prevention Study II nested case-control study | cohort/ USA | 1,086 | 533 | 548 | - | 459 | 78 | 11 | 538 | - | 457 | 76 | 5 |
| ColoCare_ Heidelberg | ColoCare Consortium | case-series/ Germany | 226 | 84 | 189 | 25 | 146 | 39 | 4 | 37 | 10 | 33 | 3 | 1 |
| ColoCare_ Seattle | ColoCare Consortium | case-series/ USA | 180 | 77 | 180 | 52 | 135 | 41 | 4 | - | - | - | - | - |
| ESTHER_VE RDI | Epidemiologische Studie zu Chancen der Verhütung, Früherkennung und optimierten Therapie chronischer Erkrankungen in der älteren Bevölkerung; Verlauf der diagnotischen Abklärung bei Krebspatienten | case-control Germany | 829 | 288 | 402 | 8 | 319 | 75 | 8 | 427 | 1 | 340 | 81 | 6 |
| Kentucky | Kentucky Case-Control Study | case-control/ USA | 2,871 | 1,432 | 1,232 | 111 | 981 | 231 | 20 | 1,639 | 9 | 1,318 | 304 | 17 |
| Kiel | PopGen Biobank | cohort/ Germany | 1,114 | 488 | 1,114 | 128 | 908 | 189 | 17 | - | - | - | - | - |

| | | | | | | | | | | | | | | |
|---|---|---|---|---|---|---|---|---|---|---|---|---|---|---|
| Korea | Korean Cancer Prevention Study-II CRC | cohort/ Korea | 5,418 | 2,505 | 3,052 | 395 | 2,388 | 616 | 48 | 2,366 | 381 | 1,900 | 431 | 35 |
| MCCS | Melbourne Collaborative Cohort Study | cohort/ Australia | 1,435 | 689 | 717 | 10 | 611 | 98 | 8 | 718 | 28 | 598 | 117 | 3 |
| MEC | Multiethnic Cohort Study | cohort/ USA | 401 | 179 | 186 | - | 157 | 25 | 4 | 215 | 11 | 181 | 30 | 4 |
| MECC | Molecular Epidemiology of Colorectal Cancer | case-control/ Israel | 7,693 | 3,706 | 4,295 | 320 | 3,034 | 1,135 | 126 | 3,398 | 162 | 2,500 | 807 | 91 |
| MSKCC | Memorial Sloan Kettering Cancer Center Cohort | case-control USA | 76 | 46 | 76 | 19 | 66 | 9 | 1 | - | - | - | - | - |
| NF | Newfoundland Case-Control Study | case-control Canada | 672 | 271 | 196 | 25 | 172 | 22 | 2 | 476 | 40 | 412 | 63 | 1 |
| NHS2 | Nurses' Health Study | cohort USA | 173 | 173 | 92 | 22 | 65 | 26 | 1 | 81 | 81 | 65 | 16 | - |
| SWEDEN_Lindblom | Swedish Low-Risk Colorectal Cancer Study (SLRCCS). | cohort/ Sweden | 4,906 | 2,267 | 2,566 | 123 | 2,211 | 334 | 21 | 2,340 | 528 | 1,998 | 329 | 13 |
| SWEDEN_ Wolk | Swedish Mammography Cohort (SMC) and the Cohort of Swedish Men (COSM), | cohort/Sweden | 1,408 | 591 | 566 | 8 | 459 | 101 | 6 | 842 | 27 | 699 | 132 | 11 |
| Shanghai | Shanghai MHS and WHS† | cohort/ Shanghai | 457 | 214 | 233 | - | 196 | 34 | 3 | 224 | - | 180 | 44 | - |
| Spain | Colorectal Cancer Genetics & Genomics, Spanish study | case-control/ Spain | 1,642 | 676 | 802 | 51 | 669 | 119 | 14 | 840 | 90 | 676 | 153 | 11 |

| | | | | | | | | | | | | | | |
|---|---|---|---|---|---|---|---|---|---|---|---|---|---|---|
| UK_SEARCH | Studies of Epidemiology and Risk Factors in Cancer Heredity | case-control UK | 4,335 | 1,857 | 4,219 | 457 | 3,563 | 622 | 34 | 116 | 21 | 97 | 19 | - |
| USC_HRT_CRC | Los Angeles County Cancer Surveillance Program | case-control/ USA | 772 | 772 | 359 | - | 291 | 66 | 2 | 413 | - | 335 | 72 | 6 |

**Supplementary Table 2. Descriptors of Validation Set Studies**

| Study | Title | Study Type/ Population | Total | Females | Cases | | | | | Controls | | | | |
|---|---|---|---|---|---|---|---|---|---|---|---|---|---|---|
| | | | | | | | Genotype | | | | | Genotype | | |
| | | | | | N | <50 | GG | GC | CC | N | <50 | GG | GC | CC |
| ASTERISK | Association STudy Evaluating RISK for sporadic colorectal cancer | case-control / France | 1838 | 763 | 892 | 33 | 8 | 142 | 742 | 946 | 65 | 16 | 164 | 766 |
| ATBC | Alpha-Tocopherol, Beta Carotene Cancer Prevention Study | cohort/ Finland | 7 | - | 4 | - | - | 3 | 1 | 3 | - | - | 2 | 1 |
| CCFR | Colon Cancer Family Registry | case-control / USA, Canada, Australia | 2173 | 1086 | 1109 | 585 | 13 | 164 | 932 | 1064 | 249 | 9 | 163 | 892 |
| CLUEII | Campaign against Cancer and Heart Disease II | cohort / USA | 531 | 285 | 272 | 46 | 5 | 49 | 218 | 259 | 43 | 2 | 51 | 206 |
| CORSA | Colorectal Cancer Study of Austria | case-control / Australia | 4383 | 1655 | 2597 | 253 | 31 | 464 | 2102 | 1786 | 350 | 23 | 332 | 1431 |
| CPSII | American Cancer Society Cancer Prevention Study II nested case-control study | cohort/ USA | 697 | 367 | 346 | 1 | 4 | 55 | 287 | 351 | 1 | 1 | 66 | 284 |
| CRCGEN | Colorectal Cancer Genetics & Genomics, Spanish study | case-control / Spain | 350 | 177 | 128 | 11 | 2 | 24 | 102 | 222 | 13 | 3 | 36 | 183 |
| Colo2&3 | Hawai'i Colorectal Cancer Studies 2&3 | case-series / USA | 211 | 94 | 87 | 10 | 1 | 14 | 72 | 124 | 12 | 1 | 20 | 103 |
| ColoCare | ColoCare Consortium | case-series / USA, Germany | 19 | 9 | 15 | 5 | - | 4 | 11 | 4 | 1 | - | 1 | 3 |
| CzechCCS | Czech Republic CCS | case-control / Czech Republic | 3297 | 1364 | 1681 | 163 | 19 | 273 | 1389 | 1616 | 770 | 10 | 255 | 1351 |
| DACHS | Darmkrebs: Chancen der Verhütung durch Screening | case-control / Germany | 6312 | 2502 | 3523 | 167 | 50 | 619 | 2854 | 2789 | 118 | 21 | 498 | 2270 |
| DALS | Diet, Activity and Lifestyle Study | case-control / USA | 2262 | 1018 | 1099 | 104 | 6 | 172 | 921 | 1163 | 106 | 8 | 180 | 975 |
| EDRN | Early Detection Research Network | case cohort / USA | 655 | 309 | 299 | 47 | 2 | 57 | 240 | 356 | 35 | 8 | 79 | 269 |
| EPIC | European Prospective Investigation into Cancer and Nutrition | cohort / Europe | 4150 | 2249 | 1926 | 354 | 22 | 309 | 1595 | 2224 | 422 | 24 | 383 | 1817 |
| EPICOLON | EPICOLON | case-control / Spain | 632 | 269 | 275 | 65 | 2 | 61 | 212 | 357 | 17 | 2 | 74 | 281 |

| | | | | | | | | | | | | | | |
|---|---|---|---|---|---|---|---|---|---|---|---|---|---|---|
| ESTHER_VERDI | Epidemiologische Studie zu Chancen der Verhütung, Früherkennung und optimierten Therapie chronischer Erkrankungen in der älteren Bevölkerung; Verlauf der diagnotischen Abklärung bei Krebspatienten | case-control/ Germany | 21 | 7 | 15 | - | 2 | 3 | 10 | 6 | - | - | 1 | 5 |
| GALEON | GALicia Estudio Oncológico de coloN | case-control / Spain | 92 | 37 | 92 | 2 | 1 | 20 | 71 | - | - | - | - | - |
| HPFS | Health Professionals Follow-Up Study | cohort / USA | 1190 | - | 588 | 13 | 6 | 110 | 472 | 602 | 13 | 12 | 101 | 489 |
| Kentucky | Kentucky Case-Control Study | case-control/ USA | 1386 | 696 | 670 | 92 | 9 | 117 | 544 | 716 | 47 | 2 | 99 | 615 |
| LCCS | Leeds Colorectal Cancer Study | case-control / UK | 2098 | 932 | 1412 | 77 | 4 | 190 | 1218 | 686 | 25 | 3 | 85 | 598 |
| MCCS | Melbourne Collaborative Cohort Study | cohort/ Australia | 1001 | 487 | 537 | 63 | 5 | 81 | 451 | 464 | 59 | 1 | 67 | 396 |
| MEC | Multiethnic Cohort Study | cohort/ USA | 1061 | 508 | 525 | 53 | 8 | 106 | 411 | 536 | 49 | 13 | 114 | 409 |
| MECC | Molecular Epidemiology of Colorectal Cancer | case-control/ Israel | 2153 | 1001 | 1030 | 75 | 44 | 298 | 688 | 1123 | 58 | 44 | 294 | 785 |
| MOFFITT | Colorectal Cancer Outcomes Prognosis and Epidemiology (COPE) Study and Total Cancer Care (TCC) | cohort / USA | 412 | 189 | 412 | 29 | 6 | 72 | 334 | - | - | - | - | - |
| NCCCSI | North Carolina Colon Cancer Study, I | case-control / USA | 1041 | 476 | 404 | 40 | 17 | 101 | 286 | 637 | 42 | 20 | 146 | 471 |
| NCCCSII | North Carolina Colon Cancer Study, II | case-control / USA | 1568 | 625 | 766 | 117 | 20 | 140 | 606 | 802 | 72 | 13 | 164 | 625 |
| NFCCR | Newfoundland Case-Control Study | case-control / Canada | 956 | 391 | 484 | 66 | 8 | 61 | 415 | 472 | 69 | - | 66 | 406 |
| NGCCS | PopGen Biobank | case-control / Germany | 1117 | 489 | 1117 | 136 | 8 | 173 | 936 | - | - | - | - | - |
| NHS | Nurses' Health Study | cohort / USA | 2098 | 2098 | 856 | 36 | 10 | 141 | 705 | 1242 | 56 | 7 | 179 | 1056 |
| NHSII | Nurses' Health Study | cohort/ USA | 43 | 43 | 20 | 10 | 1 | 4 | 15 | 23 | 10 | - | 3 | 20 |
| PHS | Physicians' Health Study | cohort / USA | 761 | - | 374 | 64 | 1 | 63 | 310 | 387 | 69 | 5 | 80 | 302 |
| PLCO | Prostate, Lung, Colorectal, and Ovarian Cancer Screening Trial | cohort / USA | 6051 | 3079 | 1863 | - | 15 | 305 | 1543 | 4188 | - | 30 | 642 | 3516 |
| PPS3 | Aspirin/Folate Polyp Prevention Study | clinical trial / USA | 531 | 205 | 73 | 8 | 1 | 17 | 55 | 458 | 91 | 6 | 81 | 371 |

| | | | | | | | | | | | | | | |
|---|---|---|---|---|---|---|---|---|---|---|---|---|---|---|
| PPS4 | Vitamin D/Calcium Polyp Prevention Study | clinical trial / USA | 238 | 107 | 32 | 1 | - | 9 | 23 | 206 | 18 | 10 | 62 | 134 |
| SEARCH | Studies of Epidemiology and Risk Factors in Cancer Heredity | case-control / UK | 1133 | 1017 | 93 | 12 | 1 | 15 | 77 | 1040 | 741 | 8 | 145 | 887 |
| SELECT | Selenium and Vitamin E Prevention Trial | clinical trial / USA | 606 | - | 303 | - | 7 | 45 | 251 | 303 | - | 5 | 60 | 238 |
| SLRCCS | Swedish Low-Risk Colorectal Cancer Study | cohort / Sweden | 4150 | 1944 | 2679 | 136 | 20 | 323 | 2336 | 1471 | 623 | 7 | 176 | 1288 |
| SMC_COSM | Swedish Mammography Cohort and Cohort of Swedish Men | cohort / Sweden | 36 | 19 | 16 | 2 | - | 1 | 15 | 20 | 2 | - | 7 | 13 |
| UKB | UK Biobank | cohort / UK | 14885 | 6274 | 2996 | 181 | 17 | 351 | 2628 | 11889 | 723 | 68 | 1489 | 10332 |
| USC_HRT_CRC | Los Angeles County Cancer Surveillance Program | case-control/ USA | 227 | 227 | 128 | - | 4 | 45 | 79 | 99 | 2 | 8 | 31 | 60 |
| VITAL | VITamins And Lifestyle | cohort / USA | 552 | 260 | 270 | - | 2 | 36 | 232 | 282 | - | - | 36 | 246 |
| WHI | Women's Health Initiative Study | cohort / USA | 4059 | 4059 | 1986 | - | 18 | 299 | 1669 | 2073 | - | 22 | 331 | 1720 |

**Supplementary Tables and Figures:**

**Supplementary Figure 1.**

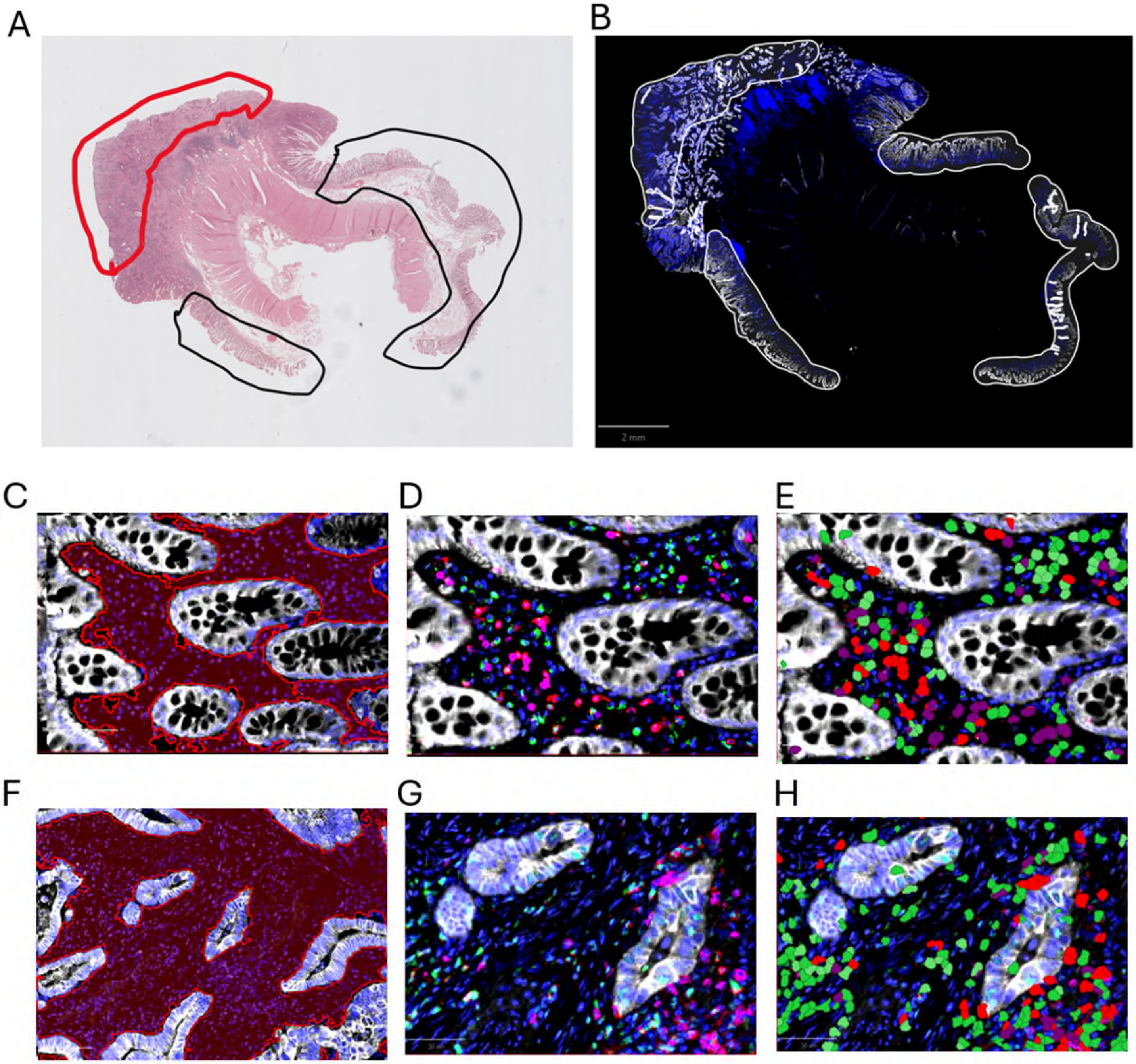

**Supplementary Figure 1: Multiplex Immunofluorescence image analysis strategy**. A. H&E images of whole slide tissues were evaluated by a pathologist to identify areas of mucosa (black circles) and carcinoma (red circles). B. Areas identified by a Board-certified pathologist were marked as regions of interest in whole slide multiplex immunofluorescence images. C. Mucosal stroma (red) was segmented from mucosal glands. D. Representative staining of CD3 (red), CD8 (magenta), CD57 (green), and pan-CK (white) in mucosal tissues. E. Representative phenotyping of CD4+ T cells (CD3+ CD8-, purple), CD8+ T cells (CD8+, red), and NK cells (CD3-, CD8-, CD57+, green). F. Tumor stroma (red) segmented from cancer islands. G. representative staining and H. Immune phenotyping in a superficial tumor area.
